# High-flow nasal cannula for acute exacerbation of chronic obstructive pulmonary disease: a Cost-utility analysis

**DOI:** 10.1101/2022.06.17.22276548

**Authors:** Jefferson Antonio Buendía, Diana Guerrero Patiño

## Abstract

**Background:** High-flow nasal cannula (HFNC) is an alternative for delivering respiratory support to adults with acute exacerbations of chronic obstructive pulmonary disease. Despite increased popularity for respiratory support, there is still uncertainty about if their l costs of justify the clinical benefits provided. This study aims to evaluate the cost-utility of HFNC in acute exacerbations of chronic obstructive pulmonary disease in Colombia

**Methods:** Using a decision tree model, we estimated the cost and quality-adjusted life-years (QALYs) associated with HFNC and conventional oxygen therapy (COT) in adults who presented to the emergency department with acute exacerbations of chronic obstructive pulmonary disease. All parameters for the model were derived from published research with local data. All analyses were done from a societal perspective.

**Results:** We estimate a gain of 0,49 and 0,48 QALYs per patient-year on HFNC and COT respectively, and a difference of US$314 in total discounted cost per person-year on HFNC respecting COT. Because HFNC was associated with lower costs compared to conventional therapy, the incremental cost effectiveness ratio was not calculated.

**Conclusions:** HFNC achieving better outcomes at a lower cost in patients with acute exacerbations of chronic obstructive pulmonary disease in Colombia. Evidence should continue to be generated with real-life effectiveness data and economic evaluations in other countries to confirm our findings.

## Introduction

Chronic obstructive pulmonary disease (COPD) is among the leading causes of morbimortality worldwide. COPD ranked eighth among the top 20 conditions causing disability globally(1). In 2015, 3.2 million people died from COPD worldwide, with an increasing trend over the last 20 years (2). Acute exacerbation of COPD is the main cause of death in these patients. Oxygen therapy with low-flow oxygen is initially the main treatment method for patients with acute exacerbation (3). However, CO2 retention appears as acute exacerbation progresses. This hypercapnia can complicate both COPD exacerbations and stable COPD (4). In some patients, this hypercapnia, with the resulting hypoxia, is difficult to correct, and non-invasive positive-pressure ventilation (NIPPV) has been proposed as the first mode of ventilation in acute respiratory failure and type II respiratory failure (5). NIPPV has been demonstrated its effectiveness to provide respiratory support in cardiogenic pulmonary edema or acute exacerbations of COPD (6). However, NIPPV prevents mobilization and oral nutrition, being poorly tolerated (7, 8). The failure of these devices to give correct respiratory support often results in the need for intubation and mechanical ventilation.

HFNC, which has been used in the neonatal setting, is a relatively new method of delivering respiratory support to adults with acute respiratory failure. With this device, a flow of up to 60 liters per minute of warmed and humidified oxygen can be delivered with few adverse reactions(9). HFNC does not need to be removed when patients talk, eat, resulting in fewer interruptions of therapy. HFNC has been associated with flushing of anatomical dead space due to high gas flow, generation of positive airway pressure, which increases functional residual capacity and improves alveolar recruitment, ability to deliver optimal humidification, leading to enhanced mucociliary transport (10). These favorable physiological effects have been reflected in the metanalysis of randomized clinical trials, with significantly lower rates of treatment failure compared and with few severe adverse events or safety issues (11). Using HFNC for COPD patients can reduce the frequency of exacerbations and improve exercise capacity and quality of life (12, 13).

A recent metanalysis of 6 randomized controlled trials with a total of 526 COPD patients shows that compared with NIPPV, HFNC can reduce the PaCO2 level, length of hospital stay, and the incidence of nasal facial skin breakdown (14). Despite increased popularity as a treatment modality for respiratory support, there is still uncertainty about if the additional costs of this device justify the clinical benefits provided. In a previous paper, we demonstrated the cost-utility of this treatment in other diseases such as acute bronchiolitis with a favorable budgetary impact on the Colombian health system (15, 16). This would be significant, especially for hospitals in middle-income countries with scarce health resources, and where this technology could be a cost-saving alternative (17). This study aims to evaluate the cost-effectiveness of HFNC in critically ill adults with hypoxemic respiratory failure.

## Methods

### Model structure

Using a decision tree analysis, we estimate the cost and quality-adjusted life-years (QALYs) associated with HFNC and conventional oxygen therapy (COT) in adults who presented to the emergency department acute exacerbations of chronic obstructive pulmonary disease (AECOPD), **figure 1**.

**Figure 1.**
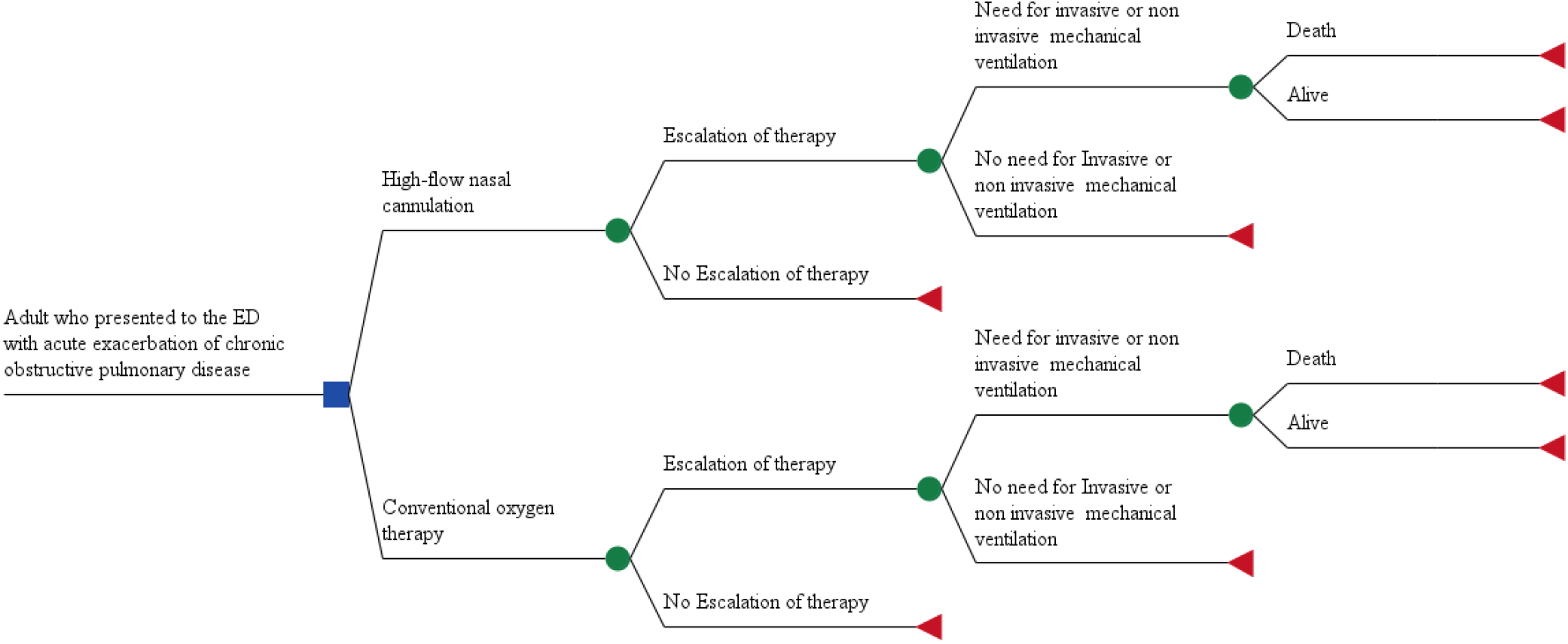
Decision tree model

The case base, and according to the inclusion and exclusion criteria of the randomized controlled trial (RCT) from which the relative risk was extracted for this mathematical model, and which will be detailed below, included patients with compensated hypercapnic respiratory failure, and who were aged 40 years or older (18). In this RCT compensated hypercapnic respiratory failure was defined as “the baseline arterial blood gas analysis (measured with room air in the supine position after at least 30 min of rest) results of pH ≥7.35, partial arterial oxygen pressure (PaO2) < 60 mmHg, and PaCO2 > 45 mmHg” (18). In our case base were excluded “patients with long-term NIPPV at home, respiratory failure requiring mechanical ventilation at admittance, isolated cardiogenic pulmonary edema indicating NIV, gastrointestinal hemorrhage, pneumothorax, or life-threatening organ dysfunction”(18). In this trial the HFNC group utilized as respiratory support high-flow devices (AIRVOTM 2; Fisher & Paykel Healthcare) and “the sizes of the nasal cannulas were chosen based on patients’ nostrils, the humidifier temperature was set to 31°C, 34°C, or 37°C according to the comfort degree of the patients, and the fraction of inspired oxygen (FiO2) was adjusted to maintain oxygen saturation by pulse oximetry (SpO2) at 90% to 93%”. The maximum flow rate also had to be adjusted according to the patients’ maximum tolerance. Patients were instructed to use HFNC for at least 15 h per day, and the total treatment time had to be no less than 5 d” (18). The group with COT receives “oxygen via nasal prongs for at least 15 h per day. Oxygen flow was set to achieve SpO2 at 90% to 93%” (18).

In our decision tree analysis, we defined the following outcomes according to the natural history of the disease: death, admission to PICU with a need for invasive or non-invasive mechanical ventilation, admission to PICU without need for invasive or non-invasive mechanical ventilation, hospitalization with the need of escalation therapy, hospitalization without the need of escalation therapy. In this analytical model, any patient who presents to the emergency department with an acute exacerbation of COPD after receiving oxygen with HFNC or COT may or may not have a requirement to escalate the therapy to invasive mechanical ventilation and after it may die or survive. In case of not requiring non-invasive mechanical ventilation (state of non-escalation of therapy), or also in the patient who survives after the requirement of mechanical ventilation, it is assumed that the patient does not have any sequelae and is discharged from the hospital, **figure 1**.

The analytic horizon was an acute episode of hypoxemic respiratory failure approximately 12 days (18). Given the short time horizon, no type of discount to costs or results was applied. Treatment was considered cost-effective if the incremental cost-utility ratio was below $5180 per QALY gained using the willingness to pay (WTP) for QALY in Colombia (19).

### Parameters of the model

All parameters for the model were derived from published research with local data, **table 1**. The relative risk of escalation of therapy and invasive mechanical ventilation of HFNC, as well the transition probabilities were derived from a prospective, randomized, controlled trial. In this RTC, 320 patients with AECOPD and compensated hypercapnic respiratory failure were recruited from general wards in a multicentric study(18). They were randomized to HFNC (with a FiO2 adjusted to obtain an oxygen saturation at 90% to 93% and with a maximum flow rate adjusted according to the patient’s tolerance) for at least 15 h per day, and the total treatment time had to be no less than 5 days) or conventional oxygen therapy or COT (oxygen delivered via nasal prongs for at least 15 h per day with a flow set to achieve SpO2 at 90% to 93%). Utilities were extracted from a prospective cohort study of 195 survivors of acute respiratory distress (20). We assumed that the lifetime QALYs were the same for all survivors, irrespective of whether they were in COT or HFNC (21).

**Table 1.** Case Base

| Variable | Base case | High Value | Low Value | Ref |
| --- | --- | --- | --- | --- |
| Cost US\$ (per /patient day) | | | | |
| High-flow nasal cannula | 58 | 72 | 43 | (23,23) |
| Escalation therapy state with need of invasive or non-invasive mechanical ventilation | 495 | 618 | 371 |  |
| Escalation therapy state without need of invasive or non-invasive mechanical ventilation | 425 | 566 | 339 |  |
| Non-escalation state | 28 | 33 | 20 |  |
| Utilities |  |  |  |  |
| Acute respiratory failure | 0,5 | 0,62 | 0,37 | (20) |
| Relative risk on probability of escalation therapy | 0,54 | 0,29 | 0,91 | (18) |
| Relative risk on probability of invasive or non-invasive mechanical ventilation | 0,52 | 0,27 | 0,94 |  |
| Transition probabilities |  |  |  |  |
| Probability of escalation therapy with conventional oxygen therapy | 0,19 | 0,4 | 0,24 | (18) |
| Probability of invasive or non-invasive mechanical ventilation in escalation therapy | 0,16 | 0,36 | 0,22 |  |
| Probability of death in patients with need of invasive or non-invasive mechanical ventilation | 0,27 | 0,34 | 0,20 |  |

All analyses were done from a societal perspective (including direct and indirect costs). All direct medical costs were incorporated into the model using the micro-costing including consultation at the emergency room, specialist referrals, chest physiotherapy, diagnosis support (laboratory, electrocardiogram, x-ray, etc.), oxygen (HFNC, invasive mechanical ventilation, COT, NIPPV) medication (corticosteroids, bronchodilators, etc.), medical devices, hotel services in the intensive care unit, hotel services and overhead cost in the general medical ward. All costs were extracted from National Drug Price Information System (SISMED, 2020), National Health Reference Price List (SOAT 2010), and local publications (22, 23). For the valuation of the indirect costs associated with parents’ loss of productivity, the human capital method was used, assuming everyone receives an income of at least the legal minimum wage for formal or informal work. The cost-opportunity of the productivity loss at the workplace and the caregiver was assessed based on the minimum wage without including transportation assistance for 2020 (US$230 per month) (24). We used US dollars (Currency rate: US$1.00 = COP$ 3,600) to express all costs in the study(25). The incremental cost-utility ratio (ICUR) was calculated using the following formulae:

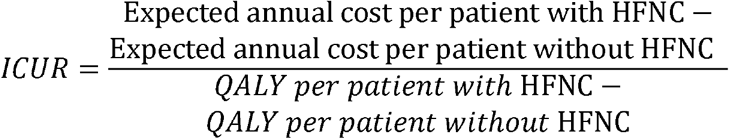

Also, we estimated the net monetary benefit (NMB). NMB represents the value of an intervention in monetary terms(26). NMB is calculated as (incremental benefit x threshold) – incremental cost. Incremental NMB measures the difference in NMB between alternative interventions, a positive incremental NMB indicating that the intervention is cost-effective compared with the alternative at the given willingness-to-pay threshold

### Sensitivity analysis

All data were subjected to probabilistic sensitivity analysis as detailed below, and as recommended by Consolidated Health Economic Evaluation Reporting Standards (CHEERS) Statement (27). To explore parameter uncertainty of the model inputs, first, we conducted a one-way sensitivity analysis represented in a tornado diagram. In this deterministic analysis, to build the range of RR to be used in this analysis, we use the CI 95% of RR mentioned before. In the case of utilities, transition probabilities, and treatment adherence, the upper and lower ranges were estimated by adding or subtracting 25% of the value from the central value defined for the base case. Then, we performed probabilistic sensitivity analysis by randomly sampling from each of the parameter distributions (beta-distribution in the case of transition probabilities, and gamma distribution in the case of costs, lognormal for length of stay, utilities, and relative risk). The expected costs and expected QALYs for each treatment strategy were calculated using that combination of parameter values in the model. This process was replicated one thousand times (i.e., second-order Monte Carlo simulation) for each treatment option resulting in the expected cost-utility. We combined a non-parametric bootstrap-based estimation of uncertainty intervals (UI 95%) and probabilistic sensitivity analysis, by drawing a vector of values from normal distributions representing the parameter uncertainties of the cost parameters, alongside 1’000 bootstrap replications(28). All analyses were done in TreeAge Pro 2021 ®.

## Results

### Case-based analysis

Base-case analyses showed that HFNC was associated with lower costs and higher QALYs than COT. The QALYs per person estimated in the model was 0,49 (UI 95% 0,41-0,50) and 0,48 (UI 95% 0,47-0,52) QALYs per patient-year on HFNC and COT, respectively. The total costs per person were US$ 965 (UI 95% 961-989) for HFNC and US$ 1271 (UI 95% 1262-1298) for COT, **table 2**. A position of dominance eliminated the need to calculate an incremental cost□utility ratio. The incremental NMB of corticosteroids plus antibiotics over without corticosteroids was of U$375 (CI 95% 360-396).

**Table 2.** Cost effectiveness analysis

| Strategy | Cost per patient (US\$) | Difference (US\$) | QUALYs | Difference | NMB(US\$) | ICUR |
| --- | --- | --- | --- | --- | --- | --- |
| HFNC | 674 |  | 0,49 |  | 8787 |  |
| COT | 988 | -314 | 0,48 | 0,01 | 8412 | (Dominated) |

NMB: Net monetary benefit
ICUR : Incremental cost-utility ratio
COT : conventional oxygen therapy
HFNC : High flow nasal cannula

In the deterministic sensitivity analysis, our base-case results were robust to variations in all assumptions and parameters except for the relative risk of HFNC, **figure 2**. If the relative risk is higher than 0,86 the ICER of HFNC would exceed the WTP of US$ 5 180, **figure 3**.

**Figure 2.**
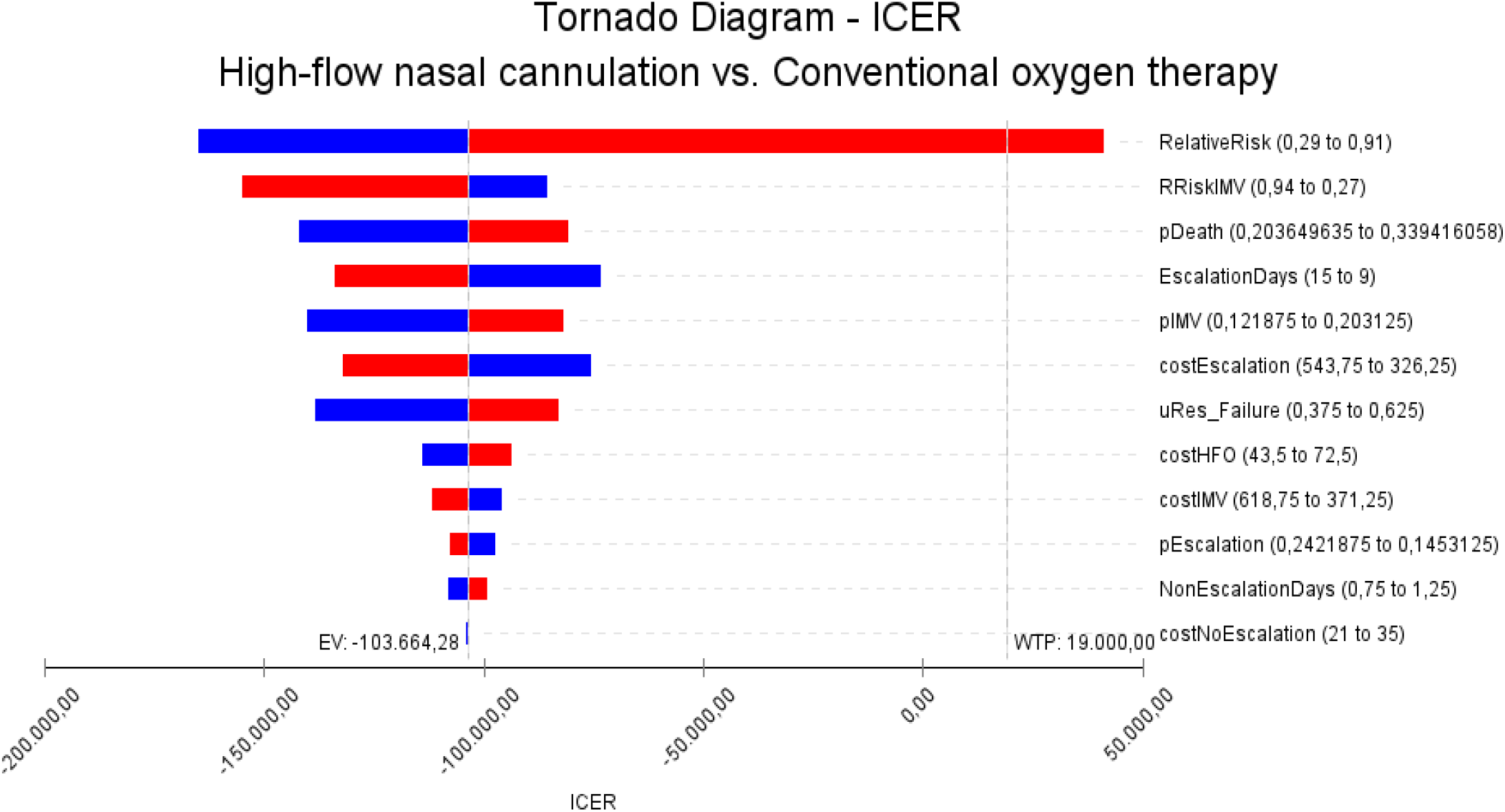
Tornado diagram

**Figure 3.**
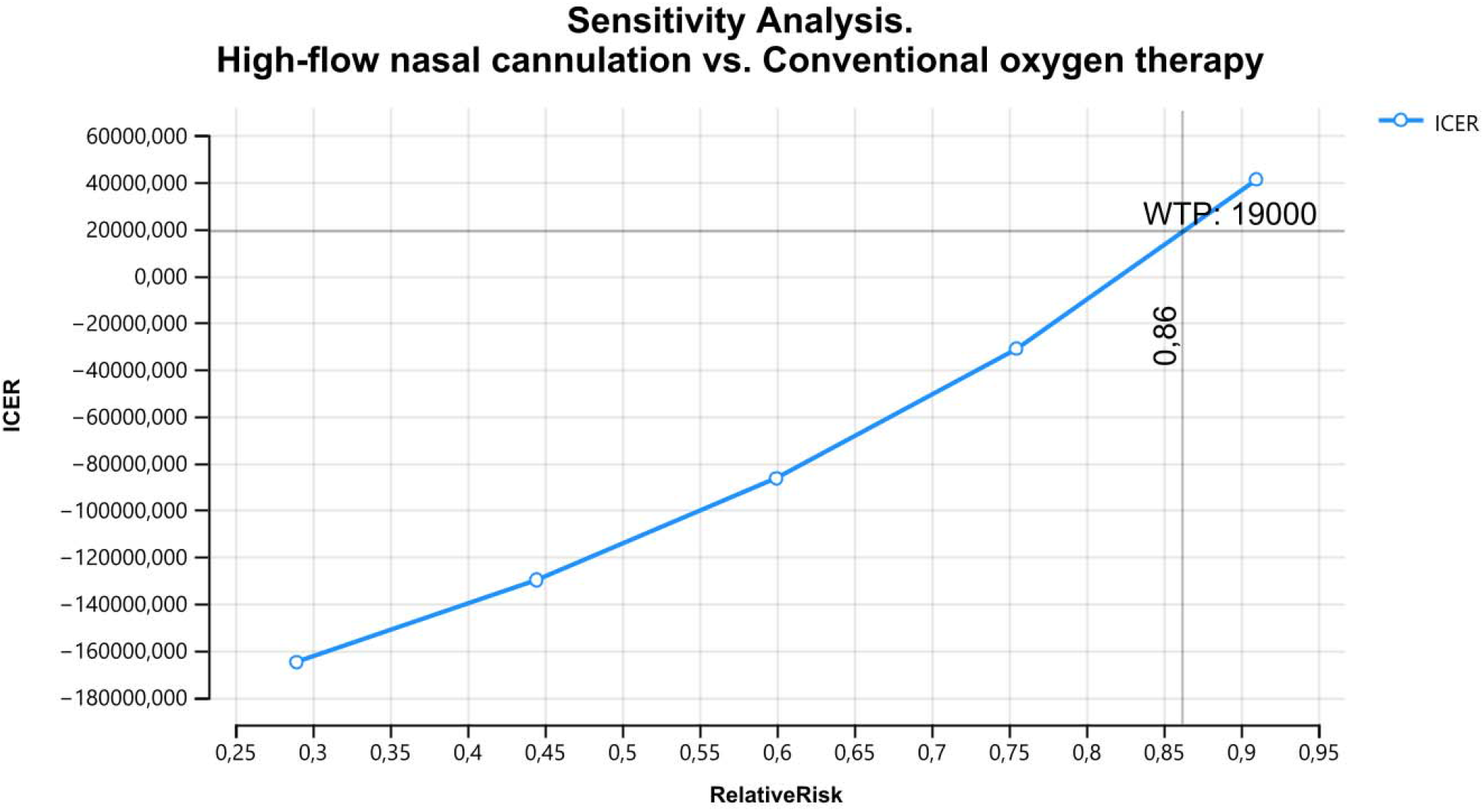
Threshold analysis of relative risk of escalation therapy of HFNC vs Conventional oxygen therapy

The results of the probabilistic sensitivity analysis are graphically represented in the cost-effectiveness plane, **figure 4**. This scatter plot shows that HFNC tends to be associated with higher costs and higher QALY. Indeed, for HFNC 86 % of ICER simulations were graphed in quadrant 2 (lower cost, high QALYs), 5% in quadrant 1 (high cost, high QALYs). The cost-effectiveness acceptability curve shows that HFNC becomes cost-effective at 100% for all willingness-to-pay thresholds, **figure 5**.

**Figure 4.**
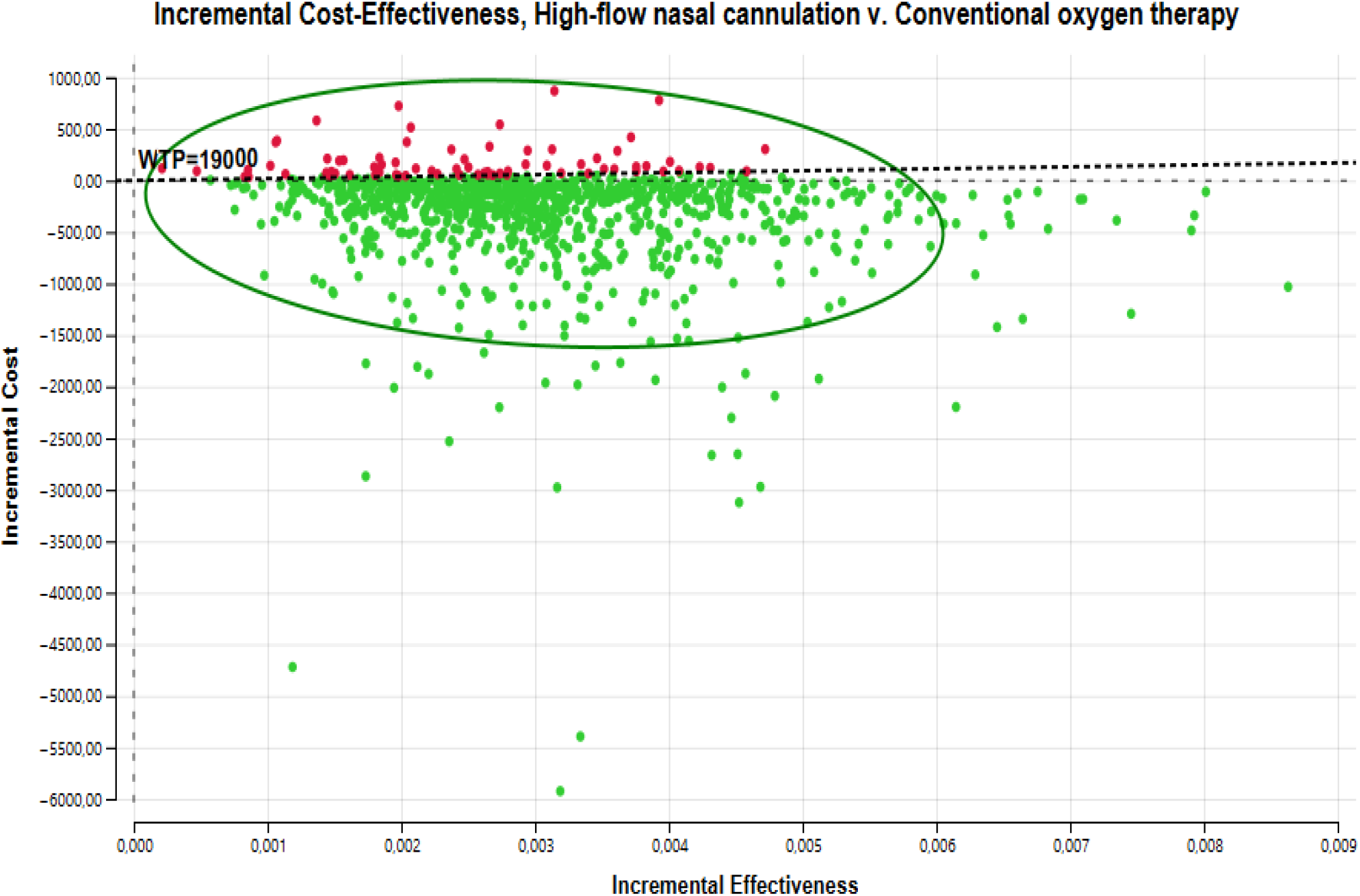
Cost effectiveness plane

**Figure 5.**
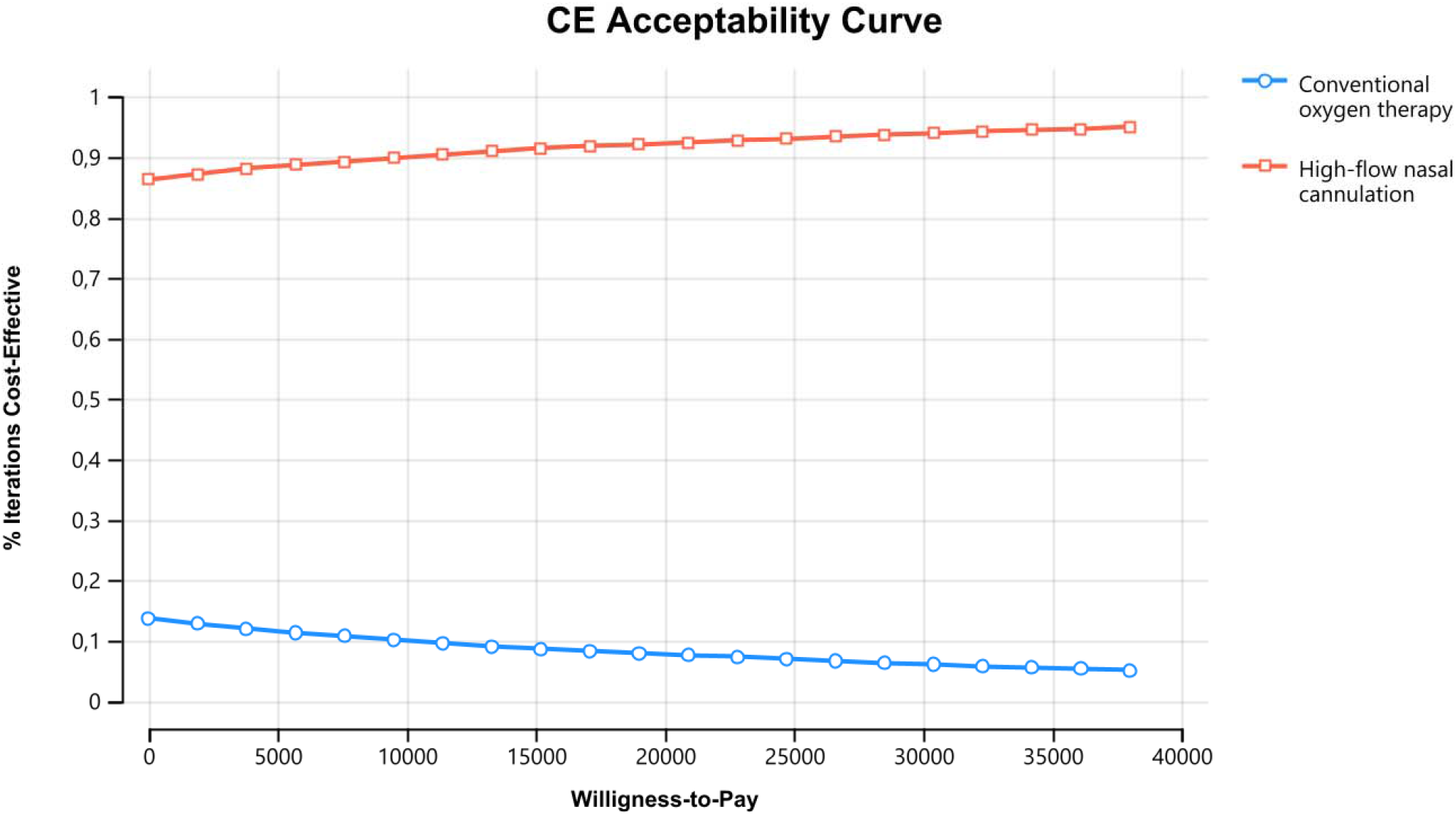
Acceptability Curve

## Discussion

Our economic analysis showed that compared with oxygen therapy by nasal catheter, HFNC, achieving better outcomes at a lower cost in AECOPD patients with acute compensated hypercapnic respiratory failure. The difference in the cost per patient between these treatments is not negligible (US$314 per patient) even more so if we consider the frequency of this disease in the population. Health economic evaluations allow translation of clinical benefits, obtained from controlled clinical trials, into composite measures of cost adjusted for quality of life-adjusted benefit units. In this case, HFNC allows for significant savings per patient, including a favorable difference regarding quality-adjusted life years. This type of evidence, although it should be replicated in other countries, allows policymakers to encourage using HFNC technologies in the clinical setting.

Our results are in line with previous evidence. In adults, with identifying a cost-effectiveness study of long-term domiciliary HFNC treatment in COPD patients with chronic respiratory failure(29). This study was done on 200 COPD patients randomized into usual care plus or without HFNC in the UK. The authors found an ICER of £3605 per QALY gained, a value below WTP in the UK (29). Turner et al, in an economic evaluation of using HFNC in intensive care units in NHS England, show that HFNC generates a cost-saving of £469 per patient compared with standard oxygen and £611 versus NIV (30). HFNC was found to dominate against standard oxygen and NIV when used in patients who had not previously been intubated and also when compared against standard oxygen in patients at low risk of re-intubation. We in a previous study estimated the cost-effectiveness of HFNC compared with oxygen by nasal cannula (control strategy) in an infant with bronchiolitis in the emergency setting(16). The cost per patient was US$368 (95% CI, US$ 323–411) in HFNC and US$441 (95% CI US$ 384–498) per patient in the control group. Also, we demonstrate in analysis the 5-year costs that high-flow nasal cannula was associated with savings for Colombian National Health equal to US$13,166,071 if the high-flow nasal cannula was adopted for the routine management of patients with acute bronchiolitis(15).

Our findings are in line with the effectiveness of HFNC reported by a recent meta-analysis. HFNC, respect to NIPPV in patients with COPD and type II respiratory failure, significantly reduce PaCO2 level (MD = − 2,64 95% CI (− 3,12 to – 2,15)), length of hospital stay ((MD = – 1,19, 95 CI (− 2,23 to – 0,05)) with a incidence of nasal facial skin breakdown ((OR = 0,11, 95% CI (0,03–0,41))(14). In patients with COPD with persistent hypercapnic and hypoxic failures, a domiciliary high-flow nasal cannula (HFNC) prevents progression in hypercapnia, the number of exacerbations, and hospitalizations (31). In this RTC 200 patients with advanced COPD and persistent hypoxic were randomized to usual care or usual care plus HFNC for 12 months. After 12 months there was a 1,3% decrease in PaCO2 in patients using HFNC and a 7% increase in controls before HFNC (p=0.003). The exacerbation rate increased, compared with 12 months; pre-study, was by 2.2/year for controls (p<0.001) and 0,15/ year for HFNC-treated patients (p=0.661). Hospital admission rates increased in the control group,+0,3/year (p=0.180), while decreasing by 0,67/year (p=0.013)for HFNC-treated patients (31). Although the evidence is still controversial insofar as there are also studies where such beneficial effects are not seen, especially in variables such as PCO2 and PO2 (32, 33), our study contributes to this knowledge by providing evidence that HFNC is cost-effective in resource-limited populations beyond intermediate discharges, given that such technology impacts the risk of escalation of therapy to more invasive and more expensive ventilation systems.

A very important aspect of our model is that it was robust to changing the model’s utility and cost values. HFNC was always the most cost-effective strategy in all ranges of cost and utilities evaluated. Although our utilities were collected from other populations, our results did not change when exploring the change in the ICER in the range of values of each utility explored. These aspects give us confidence concerning the ability to make decisions based on our results. As is always necessary for science, more studies are needed to replicate our results (34).

Our study has some limitations. As we mentioned before, we use utilities, transition probabilities, and relative risk extracted from the literature and not estimated directly from our population. As was mentioned previously, the reliability and robustness of the results were evaluated using sensitivity analysis. An additional strength is the perspective of the society on which the economic analysis was focused, which allows a faster transfer of results to health policies.

In conclusion, HFNC achieving better outcomes at a lower cost in patients with acute exacerbations of chronic obstructive pulmonary disease. Evidence should continue to be generated with real-life effectiveness data and economic evaluations in other countries to confirm our findings.

## Data Availability

All data produced in the present study are available upon reasonable request to the authors

## Acknowledgments

None

## Notes

Financial disclosures: This study was supported by own funding of authors

### Competing Interest Statement

The authors have declared no competing interest.

### Funding Statement

This study did not receive any funding

## References

1. Mortality GBD, Causes of Death C. Global, regional, and national life expectancy, all-cause mortality, and cause-specific mortality for 249 causes of death, 1980-2015: a systematic analysis for the Global Burden of Disease Study 2015. Lancet. 2016;388(10053):1459–544.

2. Collaborators GBDCRD. Global, regional, and national deaths, prevalence, disability-adjusted life years, and years lived with disability for chronic obstructive pulmonary disease and asthma, 1990-2015: a systematic analysis for the Global Burden of Disease Study 2015. Lancet Respir Med. 2017;5(9):691–706.

3. Duffy SP, Criner GJ. Chronic Obstructive Pulmonary Disease: Evaluation and Management. Med Clin North Am. 2019;103(3):453–61.

4. Steer J, Gibson GJ, Bourke SC. Predicting outcomes following hospitalization for acute exacerbations of COPD. QJM. 2010;103(11):817–29.

5. Halpin DMG, Criner GJ, Papi A, Singh D, Anzueto A, Martinez FJ, et al. Global Initiative for the Diagnosis, Management, and Prevention of Chronic Obstructive Lung Disease. The 2020 GOLD Science Committee Report on COVID-19 and Chronic Obstructive Pulmonary Disease. Am J Respir Crit Care Med. 2021;203(1):24–36.

6. Nava S, Hill N. Non-invasive ventilation in acute respiratory failure. Lancet. 2009;374(9685):250–9.

7. Bello G, De Pascale G, Antonelli M. Noninvasive Ventilation. Clin Chest Med. 2016;37(4):711–21.

8. Gregoretti C, Confalonieri M, Navalesi P, Squadrone V, Frigerio P, Beltrame F, et al. Evaluation of patient skin breakdown and comfort with a new face mask for non-invasive ventilation: a multi-center study. Intensive Care Med. 2002;28(3):278–84.

9. Nishimura M. High-flow nasal cannula oxygen therapy in adults. J Intensive Care. 2015;3(1):15.

10. Lodeserto FJ, Lettich TM, Rezaie SR. High-flow Nasal Cannula: Mechanisms of Action and Adult and Pediatric Indications. Cureus. 2018;10(11):e3639.

11. Corley A, Rickard CM, Aitken LM, Johnston A, Barnett A, Fraser JF, et al. High-flow nasal cannulae for respiratory support in adult intensive care patients. Cochrane Database Syst Rev. 2017;5:CD010172.

12. Pisani L, Astuto M, Prediletto I, Longhini F. High flow through nasal cannula in exacerbated COPD patients: a systematic review. Pulmonology. 2019;25(6):348–54.

13. Bruni A, Garofalo E, Cammarota G, Murabito P, Astuto M, Navalesi P, et al. High Flow Through Nasal Cannula in Stable and Exacerbated Chronic Obstructive Pulmonary Disease Patients. Rev Recent Clin Trials. 2019;14(4):247–60.

14. Xu Z, Zhu L, Zhan J, Liu L. The efficacy and safety of high-flow nasal cannula therapy in patients with COPD and type II respiratory failure: a meta-analysis and systematic review. Eur J Med Res. 2021;26(1):122.

15. Buendia JA, Acuna-Cordero R, Rodriguez-Martinez CE. Budget impact analysis of high-flow nasal cannula for infant bronchiolitis: the Colombian National Health System perspective. Curr Med Res Opin. 2021;37(9):1627–32.

16. Buendia JA, Acuna-Cordero R, Rodriguez-Martinez CE. The cost-utility of early use of high-flow nasal cannula in bronchiolitis. Health Econ Rev. 2021;11(1):41.

17. Slain KN, Shein SL, Rotta AT. The use of high-flow nasal cannula in the pediatric emergency department. J Pediatr (Rio J). 2017;93 Suppl 1:36–45.

18. Li XY, Tang X, Wang R, Yuan X, Zhao Y, Wang L, et al. High-Flow Nasal Cannula for Chronic Obstructive Pulmonary Disease with Acute Compensated Hypercapnic Respiratory Failure: A Randomized, Controlled Trial. Int J Chron Obstruct Pulmon Dis. 2020;15:3051–61.

19. Espinosa O, Rodriguez-Lesmes P, Orozco E, Avila D, Enriquez H, Romano G, et al. Estimating Cost-Effectiveness Thresholds Under a Managed Healthcare System: Experiences from Colombia. Health Policy Plan. 2021.

20. Ferguson ND, Scales DC, Pinto R, Wilcox ME, Cook DJ, Guyatt GH, et al. Integrating mortality and morbidity outcomes: using quality-adjusted life years in critical care trials. Am J Respir Crit Care Med. 2013;187(3):256–61.

21. Thokala P, Goodacre S, Ward M, Penn-Ashman J, Perkins GD. Cost-effectiveness of Out-of-Hospital Continuous Positive Airway Pressure for Acute Respiratory Failure. Ann Emerg Med. 2015;65(5):556–63 e6.

22. Ministerio de salud C. Sistema de Informacion de Precios de Medicamentos (SISMED) 2021 [03/07/2121]. Available from: https://www.sispro.gov.co/central-prestadores-de-servicios/Pages/SISMED-Sistema-de-Informacion-de-Precios-de-Medicamentos.aspx.

23. Enciso Oliviera C., Guerra Urrego KA, Duque M. E. MG. Costos de Atencion en UCI de un Hospital Universitario de Bogota. Repertorio de Medicina y Cirugia 2006;15(3).

24. (DANE) DNdE. Archivo nacional de datos 2019 [Available from: https://sitios.dane.gov.co/anda-index/..

25. Estadisticas DAN. Índice de Precios al Consumidor - IPC 2020 [Available from: https://www.dane.gov.co/index.php/estadisticas-por-tema/precios-y-costos/indice-de-precios-al-consumidor-ipc

26. Consortium YHE. Net Monetary Benefit [online] 2016 [Available from: https://yhec.co.uk/glossary/net-monetary-benefit/.

27. Husereau D, Drummond M, Petrou S, Carswell C, Moher D, Greenberg D, et al. Consolidated Health Economic Evaluation Reporting Standards (CHEERS) statement. Value Health. 2013;16(2):e1–5.

28. Lord J, Asante MA. Estimating uncertainty ranges for costs by the bootstrap procedure combined with probabilistic sensitivity analysis. Health Econ. 1999;8(4):323–33.

29. Sorensen SS, Storgaard LH, Weinreich UM. Cost-Effectiveness of Domiciliary High Flow Nasal Cannula Treatment in COPD Patients with Chronic Respiratory Failure. Clinicoecon Outcomes Res. 2021;13:553–64.

30. Eaton Turner E, Jenks M. Cost-effectiveness analysis of the use of high-flow oxygen through nasal cannula in intensive care units in NHS England. Expert Rev Pharmacoecon Outcomes Res. 2018;18(3):331–7.

31. Storgaard LH, Hockey HU, Weinreich UM. Development in PaCO2 over 12 months in patients with COPD with persistent hypercapnic respiratory failure treated with high-flow nasal cannula-post-hoc analysis from a randomised controlled trial. BMJ Open Respir Res. 2020;7(1). 32.

32. Huang X, Du Y, Ma Z, Zhang H, Jun L, Wang Z, et al. High-flow nasal cannula oxygen versus conventional oxygen for hypercapnic chronic obstructive pulmonary disease: A meta-analysis of randomized controlled trials. Clin Respir J. 2021;15(4):437–44.

33. Pisani L, Betti S, Biglia C, Fasano L, Catalanotti V, Prediletto I, et al. Effects of high-flow nasal cannula in patients with persistent hypercapnia after an acute COPD exacerbation: a prospective pilot study. BMC Pulm Med. 2020;20(1):12.

34. Buendia JA. [Attitudes, knowledge and beliefs of patient about anti-hypertensive drugs]. Biomedica. 2012;32(4):578–84.

